# The Omicron variant mutation at position 28,311 in the SARS-CoV-2 N gene does not perturb CDC N1 target detection

**DOI:** 10.1101/2021.12.16.21267734

**Authors:** Yanxia Bei, Kyle B. Vrtis, Janine G. Borgaro, Bradley W. Langhorst, Nicole M. Nichols

## Abstract

The emergence of new SARS-CoV-2 variants necessitates the reevaluation of current COVID-19 tests to ensure continued accuracy and reliability. The new SARS-CoV-2 variant, Omicron, is heavily mutated, with over 50 mutations within its RNA genome. Any of these mutations could adversely affect the ability of diagnostic assays to detect the virus in patient samples, potentially leading to inconclusive or false negative results. In fact, the U.S. Food and Drug Administration (FDA) has identified over two dozen diagnostic tests that contain a gene target that is expected to have “significantly reduced sensitivity due to a mutation in the SAS-CoV-2 Omicron variant”^1^. Additionally, one of the U.S. Centers for Disease Control and Prevention (CDC) Emergency Use Authorization (EUA) targets for COVID-19 tests, 2019-nCoV_N1, overlaps an Omicron mutation within the sequence targeted by the fluorescent probe. This target from the CDC has been used in many other EUA assays. Using in vitro transcribed (IVT) N gene RNA representing the wild-type (GenBank/GISAID ID MN908947.3) and Omicron variant (BA.1, GISAID ID EPI_ISL_6752027), we evaluated the performance of two different amplification protocols, both of which include the CDC 2019-nCoV_N1 primer-probe set. Both assays were able to detect the mutant N1 sequence as efficiently as the wild-type sequence. Consequently, these data suggest that diagnostic assays that use the 2019-nCoV-N1 primer-probe set are unlikely to be impacted by currently circulating Omicron lineage viruses.

## Introduction

On November 26, 2021, the World Health Organization (WHO) designated a newly identified SARS-CoV-2 variant in South Africa as a “Variant of Concern” and named it Omicron. Within days of the designation, the Omicron variant was detected in patient samples collected around the globe, including the United States. One alarming Omicron feature is the high number of mutations found within its genome, including over 30 in the spike gene, which may impact viral infectivity and antigenicity^2,3^. Early evidence indicates the Omicron variant may be more transmissible than other variants^4,5^. Furthermore, evidence from South Africa suggests the Omicron variant may be more capable of evading the immune system than the Delta variant^3,4^. However, extensive research is still required to determine how these mutations will impact the variant’s severity, transmissibility, or ability to evade the immune system.

Additionally, it is essential for researchers to determine whether these mutations decrease the effectiveness of COVID-19 diagnostic tests, many of which employ quantitative polymerase chain reaction (qPCR) assays to detect the SARS-CoV-2 genome in patient samples. In February 2020, the CDC released a qPCR-based laboratory test called the CDC 2019-Novel Coronavirus (2019-nCoV) Real-Time RT-PCR Diagnostic Panel, which targets two sites on the SARS-CoV-2 Nucleocapsid (N) gene, namely 2019-nCoV_N1 and 2019-nCoV_N2. The CDC SARS-CoV-2 assay targets have been granted EUA and have subsequently been incorporated in various SARS-CoV-2 diagnostic assays. The Omicron variant carries a C to U mutation at genomic position 28,311, which corresponds to the 3^rd^ nucleotide from the 5’ end of the 2019-nCoV_N1 probe target sequence (Figures 1A and 1B). It is important to evaluate how this mutation at 28,311 impacts N1 target detection to avoid potential false negative results from samples with the Omicron variant. Herein, we compare qPCR amplification efficacy for the CDC N1 target in wild-type and Omicron variant N gene synthetic RNA sequences using two different amplification protocols, and we find amplification to be unperturbed by the Omicron mutation at position 28,311.

**Figure 1.**
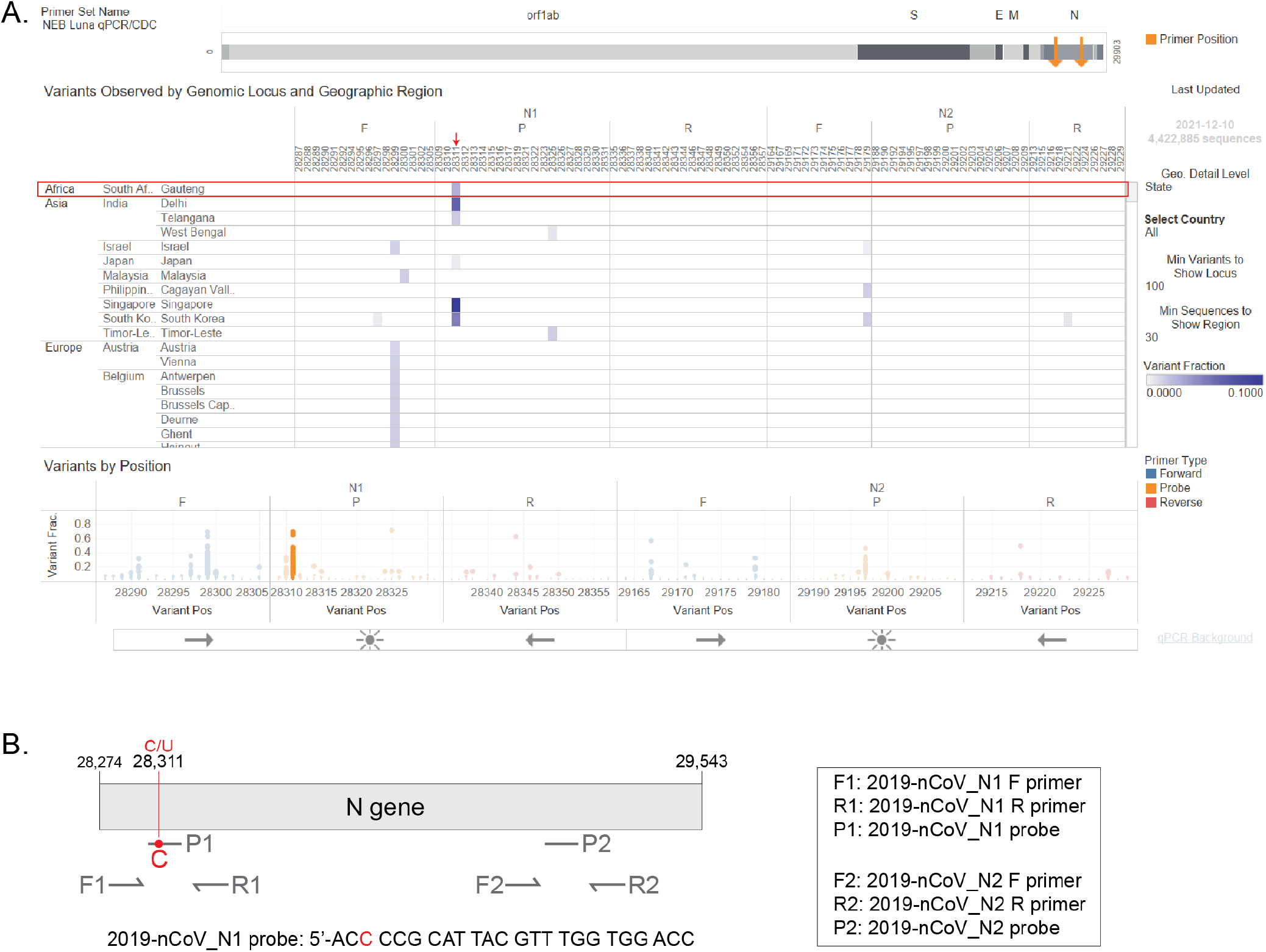
The Omicron N gene contains a mutation targeted by the 2019-nCoV_N1 detection probe. The CDC 2019-Novel Coronavirus (2019-nCoV) Real-Time RT-PCR Diagnostic Panel includes two primer-probe sets that target the SARS-CoV-2 N gene, named 2019-nCoV_N1 and 2019-nCoV_N2. A) Visual depiction of Omicron variant mutation within the N1 probe site generated using the Primer Monitor online tool (Tableau worksheet exported from Primer Monitor tool, primer set name “NEB Luna qPCR/CDC”). The Omicron variant from Gauteng, South Africa and the mutation are annotated with a red box and an arrow, respectively. Importantly, only one mutation overlapping with the N1 probe region was identified as of 12/10/2021. A second mutation in this region was identified during the preparation of this manuscript (Supplemental Figure 2A). B) Schematic representation of the two CDC primer-probe sets. Each set includes one forward primer (F), one reverse primer (R), and one fluorescent probe (P). The SARS-CoV-2 Omicron variant has a C to U mutation at position 28,311, which is within the 2019-nCoV_N1 probe (P1) target sequence. Not drawn to scale.

## Results

### NEB SARS-CoV-2 Multiplex Assay efficiently detects the N1 target from N gene RNA containing the Omicron variant mutation

The Omicron variant contains a C to U mutation at position 28,311, which corresponds to the 3^rd^ nucleotide position within the CDC 2019-nCoV_N1 probe target sequence. Given the location of this mismatch, it is unlikely that the CDC 2019-nCoV_N1 target would fail completely, but impacts to assay sensitivity cannot be dismissed. In order to test amplification of this variant RNA, we generated mutant and wild-type N gene RNA templates by IVT. Correct size and high-purity were confirmed via agarose gel (Supplemental Fig. 1). We then compared amplification of the mutant RNA to the wild-type N gene RNA using the NEB SARS-CoV-2 RT-qPCR Multiplex Assay Kit (E3019). This assay kit simultaneously detects the N1 (HEX), N2 (FAM) and the human RNase P (Cy5) targets. The N1 and N2 targets were amplified efficiently across a 7-log dilution (10^7^-10 copies/reaction) of the mutant and wild-type input RNA (Figures 2A and 2B). In these experiments, the N2 primer-probe set can also serve as an internal control to correct minor differences in RNA template input because the Omicron variant does not have a mutation within the region targeted by the N2 primers or the probe (Fig 1). The mutant RNA crossed threshold ∼1 cycle faster than wild-type RNA, likely due to slightly higher input amount. After correcting the RNA input amount based on the N2 target, there is less than a 0.2 difference in the average ΔC_q_ for the N1 target amplification between the mutant and wild-type RNA (Figures 2C and 2D). This is well within normal day-to-day and user-to-user variation and suggests equivalence in amplification speed. Importantly, the mutation did not decrease assay sensitivity as 27 out of 27 reactions with low RNA copy number (10 copies per reaction) were detected for both the wild-type and mutant RNA with both the N1 and N2 targets (Figure 2E). These data suggest the NEB SARS-CoV-2 RT-qPCR Assay can efficiently detect the IVT Omicron N gene using the CDC 2019-nCoV_N1 primer-probe set.

**Figure 2.**
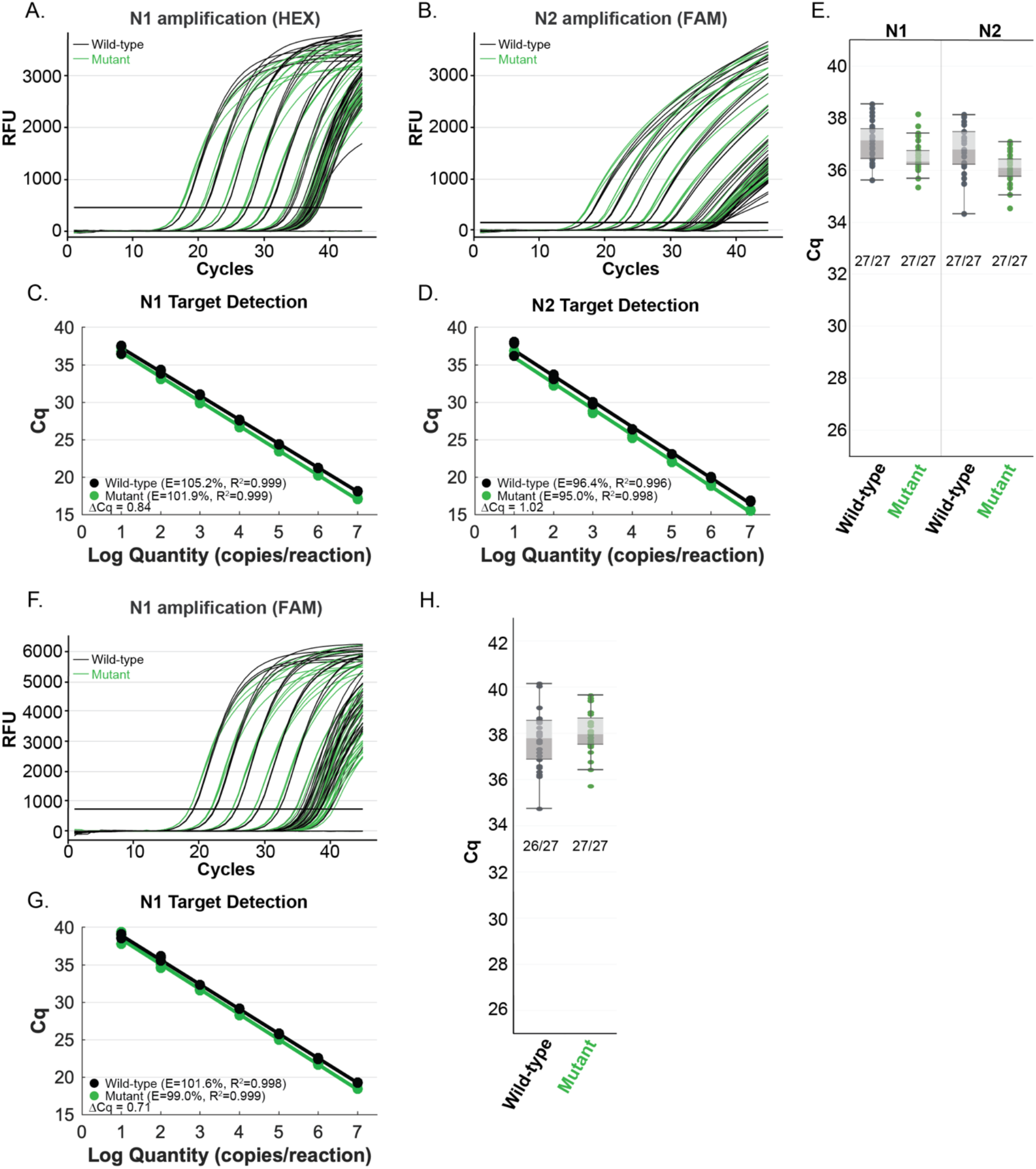
The NEB SARS-CoV-2 Multiplex Assay (A-E) and the SalivaDirect RT-qPCR protocol (F-H) efficiently detect N gene IVT RNA carrying the Omicron mutation with the CDC 2019-nCoV_N1 primer-probe set. Amplification efficiency in A-D and F-G was evaluated in triplicate over a 7-log range (10^7^-10 copies/reaction) of synthetic wild-type RNA (black) versus the Omicron mutant RNA (green). Detection sensitivity (E and H) was evaluated with 10 copies of RNA per reaction with 27 replicates per condition. All reactions tested with 10 copies of input RNA were detected by qPCR, except one wild-type sample amplified using the SalivaDirect RT-qPCR protocol.

### N gene RNA containing the Omicron variant mutation can be detected using SalivaDirect qPCR amplification conditions

SalivaDirect is a simplified, non-invasive and flexible SARS-CoV-2 diagnostic platform granted EUA by the FDA^6^. It is a two-plex assay that leverages the CDC 2019-nCoV_N1 target and human RNase P gene (RP) as an internal control. Using the RT-qPCR protocol as specified in the SalivaDirect assay, we observed efficient amplification of both the mutant and wild-type RNA across a 7-log dilution series of RNA (Figure 2F). The mutant RNA was detected slightly faster with an average ΔC_q_ of 0.71 (Figure 2G). Since the same RNA dilution series were used in both the SalivaDirect assay and the Luna SARS-CoV-2 RT-qPCR Assay (described above), the slightly faster mutant RNA detection was likely due to a higher input amount of the mutant RNA. Using purified RNA as input material, this assay was also highly sensitive for the mutant N gene, as 10 copies of input RNA were detected in 27 out of 27 samples (Figure 2H). Consequently, we conclude that both the NEB SARS-CoV-2 Multiplex Assay Kit and the SalivaDirect amplification conditions can reliably detect the Omicron N gene using the CDC 2019-nCoV_N1 primer-probe set.

## Discussion

The heavily mutated Omicron variant is concerning for many reasons, including whether the mutations could decrease the efficacy of COVID-19 diagnostic tests. The Omicron N gene harbors a mutation corresponding to the CDC-designed N1 target probe sequence in the 2019-nCoV panel, which has been incorporated into many diagnostic assays. Fortunately, our evaluation of the performances of two different amplification conditions found that the N1 site is efficiently detected with the CDC 2019-nCoV_N1 primer-probe set, despite the target probe sequence mutation. Although our evaluation has determined that the mutation does not diminish amplification performance using the CDC 2019-nCoV_N1 primer-probe set in our optimized conditions using purified RNA, additional research is required to determine whether other assay workflows or targets could be impacted.

As more variants emerge, it is essential that researchers continuously track new mutations and assess their potential impact on diagnostic assays. The Primer Monitor tool (https://primer-monitor.neb.com/) can assist in identifying potentially problematic mutations by continually monitoring registered primer-probe sets for overlapping variant mutations^7^. As previously noted, the tool revealed an overlap between an Omicron mutation at position 28,311 and the CDC 2019-nCoV_N1 probe target sequence (Figure 1A). We have also used the Primer Monitor tool to evaluate other mutations in SARS-CoV-2 variants overlapping with the CDC 2019-nCoV panel. After testing, some of these mutations decreased assay sensitivity^7^, whereas others, such as the AY43 (Delta) variant, did not impact N gene assay sensitivity (Supplemental Figure 2). During preparation of this manuscript, the Primer Monitor tool helped us identify another variant circulating in Gauteng, South Africa with a genetic mutation at position 28,320, which is also within the CDC 2019-nCoV_N1 probe target sequence (Supplemental Figure 3). This variant mutation is not shared with any published Omicron sequences. Additional testing will be required to determine the impact of this mutation on probe detection. The CDC has designed an alternative multiplex assay that can simultaneously detect genetic material from SARS-CoV-2, influenza A, and influenza B (Flu SC2 Multiplex Assay). Both CDC tests detect fragments of the N gene, however, the CDC Flu SC2 Multiplex Assay uses a different primer-probe set to detect the N gene. Currently, there are no published Omicron variant mutations within this primer-probe set. Tracking the emergence of new variant mutations and testing the performance of primer-probe sets with the variant sequences by qPCR will help ensure the continued reliability of COVID-19 diagnostic tests.

## Methods

To generate the SARS-CoV-2 N gene RNA harboring the C to U mutation at position 28,311, site-directed mutagenesis (Q5 Site-Directed Mutagenesis Kit, NEB E0554S) was performed using the SARS-CoV-2 Positive Control (N gene) plasmid (NEB N2117) as a template, and the mutation was confirmed by Sanger Sequencing. The wild-type and mutant N gene RNA was subsequently synthesized by in vitro transcription (HiScribe T7 High Yield Synthesis Kit, NEB E2040) from linearized plasmids containing either wild-type or mutant N genes, respectively. The resulting RNA was purified using the Monarch RNA Cleanup Kit (NEB, E2050) and quantitated with the Qubit RNA BR Assay Kit (ThermoFisher Scientific, Q33224) to calculate the RNA copy number. The purity and quality of the plasmids and RNA were assessed on a 1.2% agarose gel, electrophoresed for 1 h in 0.5x Tris-Borate-EDTA Buffer.

To evaluate the impact of the 28,311 C to U mutation on the 2019-nCov_N1 target detection, a 7-log dilution series (10^7^-10 copies/reaction) was prepared for both the wild-type and the mutant RNA, with 10 ng of Jurkat total RNA (BioChain, R1255815-50) included as an internal control. The mutant and wild-type target RNA were subsequently amplified using either the NEB Luna SARS-CoV-2 RT-qPCR Multiplex Assay Kit (E3019) or the Luna Universal Probe One-Step RT-qPCR Kit (NEB E3006) following the SalivaDirect RT-qPCR amplification protocol^6^. For sensitivity evaluations, 27 reactions containing 10 copies/reaction were performed using either wild-type or mutant RNA. Briefly, for the Luna SARS-CoV-2 RT-qPCR assay, the N1 (HEX), N2 (FAM) and RP (Cy5) targets were simultaneously detected using the following cycling conditions: carryover prevention (25°C for 30 s), cDNA synthesis (55°C for 10 min), initial denaturation (95°C for 1 min) and 45 cycles of denaturation (95°C for 10 sec) and annealing/elongation (60°C for 30 sec) plus a plate read step. For the SalivaDirect RT-qPCR, the Luna Universal Probe One-Step RT-qPCR Kit (NEB E3006) was used to detect the N1 (FAM) and the RP (Cy5) targets simultaneously using the following cycling conditions: cDNA synthesis step (52°C for 10 min), initial denaturation (95°C for 2 min) and 45 cycles of denaturation (95°C for 10 sec) and annealing/elongation (55°C for 30 sec) plus a plate read step. The qPCR data was collected on a Bio-Rad CFX96 qPCR instrument (96-well, 20 µl reactions).

## Data Availability

All data produced in the present study are available upon reasonable request to the authors

## Acknowledgements

We thank the NEB DNA Sequencing Core for Sanger sequencing. We also acknowledge Matt Campbell for assistance with the Primer Monitor tool.

