## Supplemental Information for "The Omicron variant mutation at position 28,311 in the SARS-CoV-2 N gene does not perturb CDC N1 target detection"

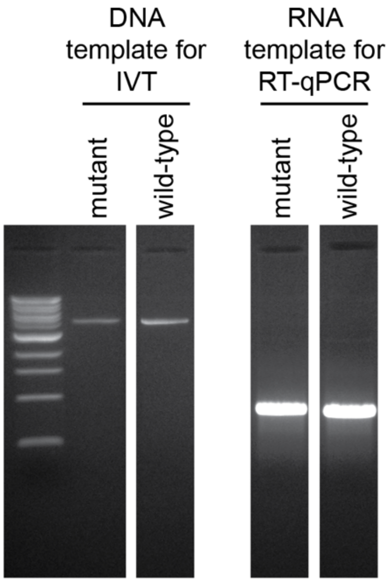


**Supplemental Fig. 1.** The quality of the linearized DNA plasmids used as templates for in vitro transcription (IVT) and the RNA yielded from IVT were assessed on a 1.2% agarose gel, ran for 1 h in 0.5x Tris-Borate-EDTA Buffer. Irrelevant lanes between the panels were removed.

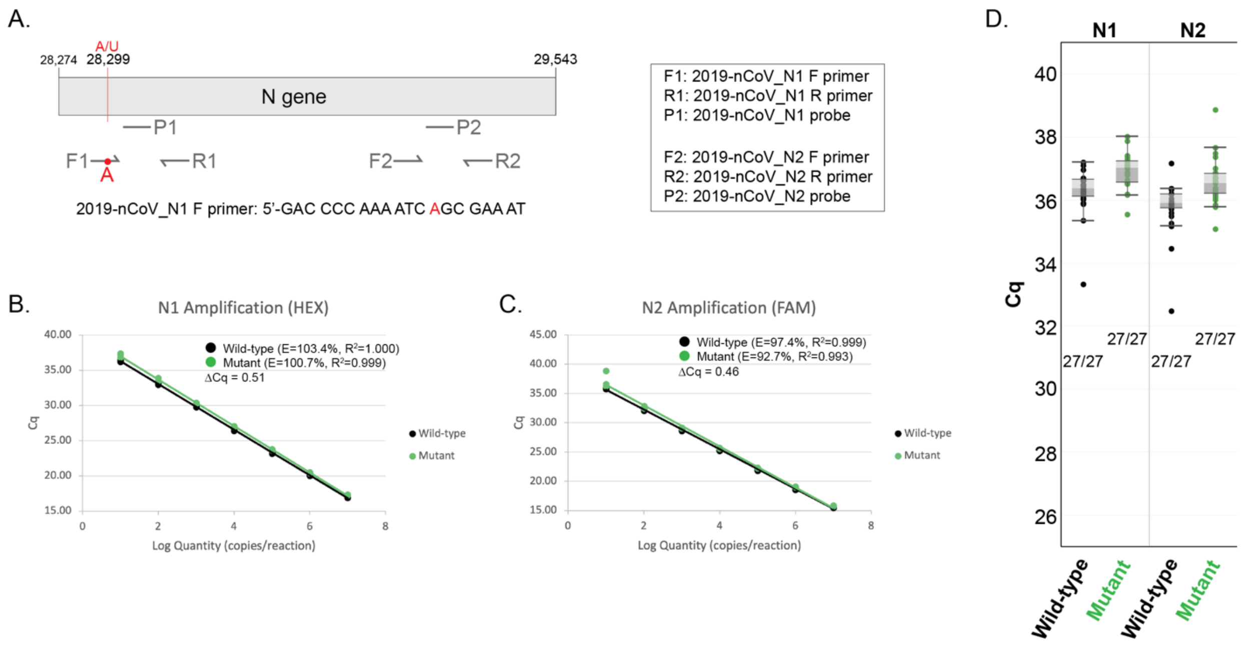


**Supplemental Fig. 2. The NEB SARS-CoV-2 Multiplex Assay efficiently detects N gene IVT RNA carrying the AY43 (Delta) variant mutation with the CDC 2019-nCoV_N1 primer-probe set**. A) Schematic representation of the two CDC primer-probe sets. Each set includes one forward primer (F), one reverse primer (R), and one fluorescent probe (P). The SARS-CoV-2 AY43 (Delta) variant has an A to U mutation (red) at position 28,299, which overlaps with the 2019-nCoV_N1 forward primer (F1) target sequence. Not drawn to scale. Amplification efficiency was evaluated at the N1 (B) and N2 (C) target sites in triplicate over a 7-log range ($10^7$-10 copies/reaction) of IVT wild-type RNA (black) versus the IVT AY43 mutant RNA (green). D) Detection sensitivity was evaluated with 10 copies of RNA per reaction with 27 replicates per condition. All reactions tested with 10 copies of input RNA were detected by qPCR.

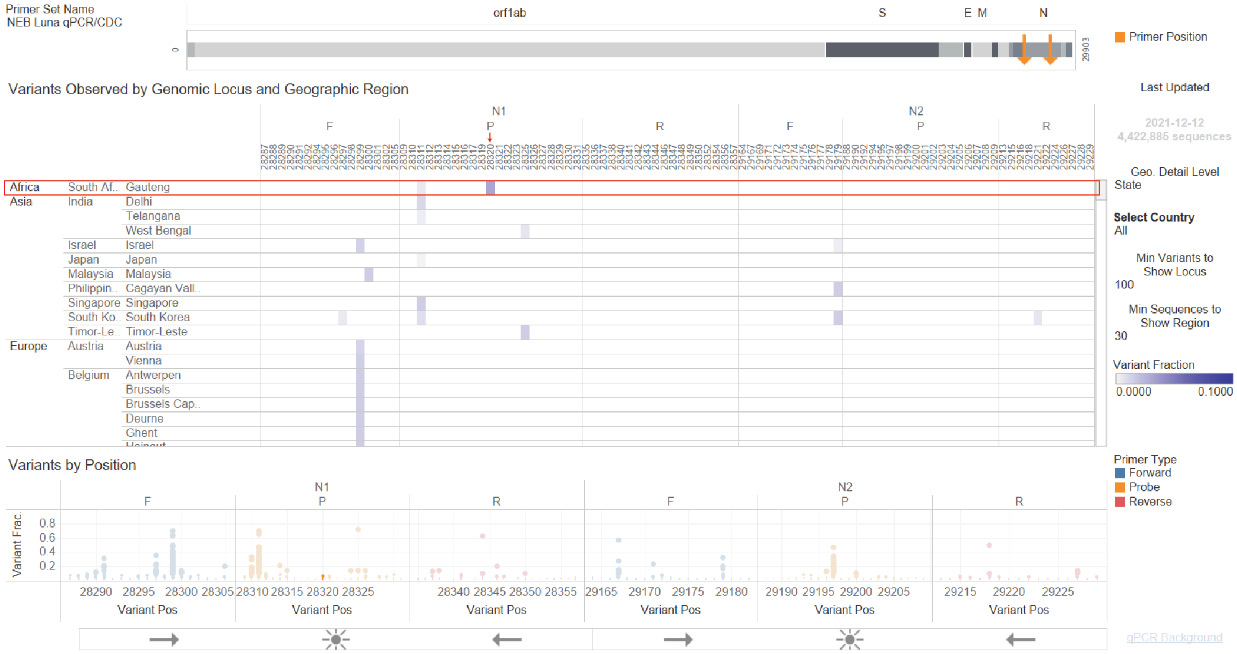


**Supplemental Fig. 3. Second mutation at genetic position 28,320 overlaps with CDC 2019-nCoV probe target site.** The mutation at genetic position 28,320 was identified using the Primer Monitor tool on 12/13/2021. The variants from Gauteng, South Africa and the mutation at position 28,320 are annotated with a red box and an arrow, respectively.
